# Female Sex Workers and Cervical Cancer Screening in Kilimanjaro Region: Uptake and Behavioural Determinants. A Community-Based Cross-Sectional Study Using the Health Belief Model

**DOI:** 10.1101/2025.05.01.25326792

**Authors:** Gumbo D. Silas, Innocent H. Uggh, Bernard Njau, Esther Majaliwa, Patricia Swai, Alma Redson, Gaudensia Olomi, John E. Mtenga, Prisca Marandu, Leah Mmari, Happiness Kilamwai, Bariki Mchome, Blandina T. Mmbaga, Alex Mremi

## Abstract

**Objective:** Cervical cancer is the 4th most leading cause of cancer-related deaths among women globally. Despite its significance, the uptake and behavioural determinants of screening among female sex workers (FSWs) in Tanzania remain poorly understood. This study aimed to address this gap among this risky population.

**Design:** A community-based cross-sectional study design.

**Setting:** This hotspot-based study was conducted at the community level, in two councils within Kilimanjaro Region, Tanzania.

**Participants:** This study involved 351 FSWs aged 25-49 years, enrolled between May and December 2024, with no history of total hysterectomy. Data was collected using an interviewer-administered questionnaire using the Respondent-Driven Sampling (RDS) technique with 0.75-0.92 Cronbach’s Alpha values for internal consistency of item questions. Analysis was carried out using SPSS version 27.0. Binary logistic regression was performed, with variables having a p-value less than 0.05 in multivariate analysis considered statistically significant.

**Main outcome measures:** Cervical cancer screening uptake.

**Results:** Out of 351 FSWs enrolled, the mean age was (36.11±5.24) years, with most living in urban areas 232(66.1%) and having completed primary education 183(52.1%). Only 17 (4.8%) had ever been screened for cervical cancer in their lifetime with the highest screening rates observed among those aged 45-49 years 3(12.5%), and those residing in urban areas 13(5.6%).

After adjusting for the modifying factors, perceived severity (AOR: 3.25, 95%CI: 1.16-9.07), perceived benefits (AOR: 3.61, 95%CI: 1.10-11.84), self-efficacy (AOR: 3.59, 95%CI: 1.18-10.96), and cues to action (AOR:3.61, 95%CI:1.28-10.15) were significantly associated with the uptake of cervical cancer screening among this population.

**Conclusion:** The uptake of cervical cancer screening among FSWs in Kilimanjaro region was extremely low. To address this challenge, targeted interventions that address key behavioural determinants, such as perceived severity, perceived benefits, self-efficacy, and cues to action, are highly recommended for this population.

**Article Summary:** *Strengths and Limitations of this study:*

- This was the first study in Tanzania focusing on FSWs, a high-risk and often underserved population.
- This study serves as a baseline for future studies and offers critical data to inform national policies, public health strategies and programs for cervical cancer prevention tailored to vulnerable groups.
- The use of the Health Belief Model (HBM) in this study, provides a structured understanding of behavioural determinants of cervical cancer screening uptake among this population.
- The use of Respondent-driven sampling (RDS) techniques in this study reduces sampling bias and provides robust data, thereby enhancing the reliability of the findings.
- However, the reliance on self-reported information for certain aspects of this study may have impacted on the accuracy of our findings due to potential recall and social desirability biases. Therefore, future studies should consider using more objective measures to minimize these biases.

## INTRODUCTION

### Background

Globally cervical cancer is ranked to be the fourth most prevalent cancer among women with approximately 661,021 new cases and 350,000 deaths recorded in 2022, accounting for 14.1% and 7.1% of all women cancer cases and deaths respectively. [1] Almost 94% of all global cervical cancer cases are from low and middle-income countries whereby in the top 20 countries with the high burden worldwide 18 are from sub-Saharan Africa, which account for 23% of global mortality rates attributable to cervical cancer[2].

Cervical cancer poses a significant health burden in the East African region, with about 40 new cases and 28.6 deaths per 100,000 women-years annually [3]. Tanzania has the highest rates within the region, reporting 59.1 new cases and 42.7 deaths per 100,000 women-years as of 2018 [3].

Female sex workers (FSWs) are highly vulnerable due to their frequent engagement in high-risk sexual behaviors, such as having multiple sexual partners, inconsistent condom use, and incentives for unsafe sexual practices [4]. These behaviors increase their risk of contracting sexually transmitted infections, including human papillomavirus (HPV), a major precursor of cervical cancer. However, HPV vaccination, regular cervical cancer screening, and treatment of precancerous lesions are globally recognized as the most effective strategies for cervical cancer prevention [5].

In 2020, the World Health Organization (WHO) launched a global strategy to accelerate the elimination of cervical cancer as a public health challenge, with three ambitious targets to be achieved by 2030 [5]. The strategy aims for at least 90% HPV vaccination coverage among adolescent girls aged 9-14 years by age 15, at least 70% cervical cancer screening coverage among immunocompetent women aged 30-49 years, and immunocompromised women aged 25-49 years using a high-performance screening test at least twice in their lifetime, and at least 90% treatment of women diagnosed with precancerous lesions and suitable management for 90% of those diagnosed with invasive cancer [5]. This screening strategy focuses on early detection of precancerous lesions, allowing for early intervention and improved outcomes[5].

Global, cervical cancer screening coverage stands at approximately 36% [2], with High-income countries achieving higher rates at 84%, compared to low-income countries that lag far behind at just 9%[6]. In sub-Saharan Africa, the coverage is only 14%[6], and in the East African region, it is slightly lower at 13% [7].

Cervical cancer screening uptake among FSWs stands at approximately 24%, globally. [8]. In Sub-Saharan Africa, the uptake appears to be lower, with a study in Ethiopia showing that only 20.3% of FSWs have ever been screened for cervical cancer.[9]. This suggests that while global screening rates among FSWs are already suboptimal, the situation is even more concerning in Sub-Saharan Africa, highlighting the need for targeted interventions to improve access to and uptake of cervical cancer screening among this high-risk population.

Tanzania adopted the global strategy for the elimination of cervical cancer as a public health problem in 2018, whereby cervical cancer screening among eligible women started. [3]. Despite these efforts to enhance cervical cancer screening in Tanzania, the uptake remains critically low, with only 7% of eligible women participating in screening [10], far below the national target of 60% by 2025. Additionally, disparities exist among subpopulations, with FSWs receiving less attention despite their heightened vulnerability to cervical cancer [11].

In the context of cervical cancer screening uptake while employing the HBM model which suggests that women are more likely to participate in screening if they perceive themselves to be at risk for cervical cancer (perceived susceptibility), recognize the severity of the disease as life-threatening (perceived severity), and believe that undergoing screening offers significant benefits, such as early detection and treatment, compared to any perceived obstacles like cost, fear, or lack of access (perceived barriers).

Additionally, their likelihood of acting is increased if they feel confident in their ability to overcome these barriers (self-efficacy). Triggers such as health campaigns, recommendations from healthcare providers, or personal experiences (cues to action) can further motivate them to undergo screening.

Therefore, understanding these behavioural determinants of cervical cancer screening among FSWs will guide the development of targeted interventions tailored to the needs of this high-risk population that is often underserved and inform public health policy and strategies that advocate for equity in the accessibility of healthcare services. From a Health Belief Model perspective, this study aimed to investigate the uptake and behavioural determinants of cervical cancer screening among FSWs in Kilimanjaro region.

## THEORETICAL FRAMEWORK

**Figure 1:**
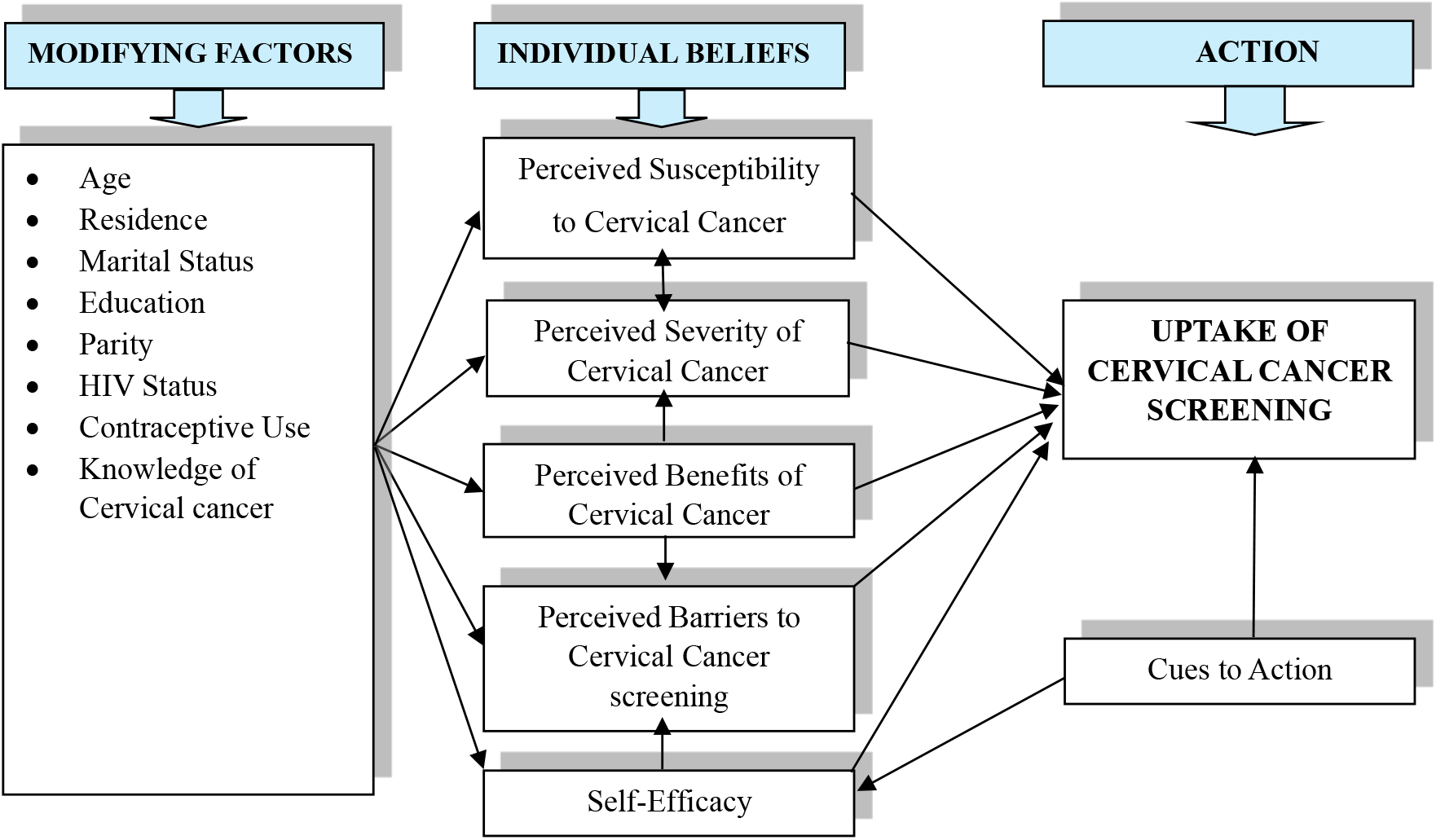
Theoretical Framework in Health Belief Model Perspective[26]. Theoretical Framework in Health Belief Model (HBM): The six constructs; Perceived susceptibility to cervical cancer, perceived severity of cervical cancer, perceived benefits of screening, perceived barriers to screening, self-efficacy, and Cues to actions represent an individual’s beliefs regarding cervical cancer and its prevention. These beliefs will determine an individual’s (FSWs) decision to undergo cervical cancer screening.[12]

## METHODS

### Study Design

This was a community-based cross-sectional study.

### Study Setting

This study was conducted in Moshi Municipal and Moshi District councils in Kilimanjaro region, Northern Tanzania, from May to December 2024.

According to the 2022 National Population and Housing Census, the region has a population of 1,861,934 (907,636 males and 954,298 females)[13]. It is bordered to the north and east by Kenya, to the south by the Tanga region, to the southwest by the Manyara region, and the west by the Manyara region. Administratively, the region is divided into seven councils: Moshi District, Moshi Municipality, Hai District, Siha District, Rombo District, Mwanga District, and Same District, with the Chaga and Pare being the dominant ethnic groups.

### Participants

This study was conducted among female sex workers (FSWs) aged 25-49 years, living with or without HIV, who engaged in transactional sex as their primary source of income for at least six months, with no history of undergoing total hysterectomy.

### Sample size and Sampling technique

Two councils, Moshi District and Moshi Municipality were purposively selected due to their high concentration of FSWs driven by the nature of economic activities that tend to attract them. Guided by a hotspot mapping checklist, a total of 12 hotspots were identified, 5 from Moshi district and 7 from Moshi municipality. A respondent-driven sampling (RDS) technique was used to enroll participants. The sample size of 351, was initially determined using a single proportion formula below:

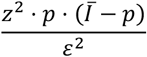

Where: **Z**: z-score for 95% confidence level, **P**: Proportion of FSWs screened for cervical cancer, **1-P**: proportion of FSWs who had never been screened, and **?**: Margin of error or precision level.

### Data Collection Process

The data collection process began with recruiting four trained peer educators (seeds), followed by identifying the network size for each seed. In wave 1, each seed was tasked with recruiting four participants through coupon distribution. In wave 2, each newly recruited participant was similarly tasked with recruiting four additional participants. The process persisted until saturation was reached.

### Variables

The uptake of cervical cancer screening, with binary response categories, was the primary outcome of this study. It was defined as having undergone a screening test of any modality in one’s lifetime. Behavioural determinants assessed using the Health Belief Model constructs served as the main exposures while other factors including sociodemographic factors were considered potential confounders or effect modifiers and were adjusted for in the analysis.

### Tool and Data source

A structured, interviewer-administered questionnaire, with Likert, scaled Item questions rated 1 to 5 adapted from similar previous studies was used. The tool was initially modified to fit the study objectives then translated from English to Swahili language and pretested among 5% of the sample size not included in the main study was used. The internal consistency of the scale used was assessed by Cronbach’s Alpha, giving values ranging from 0.75 to 0.92. Data on sociodemographic information and uptake of cervical cancer screening were self-reported while HIV status was confirmed through testing following participants’ consent.

### Data analysis

SPSS version 27.0 was used. In descriptive statistics, categorical variables were summarized in frequencies and percentages, and continuous variables in means and standard deviations. In inferential statistics, the Chi-square test assessed associations between categorical predictor variables and the outcome variable. Binary logistic regression was performed to determine the strength of the associations. All variables of a p-value less than 0.1 in the bivariate analysis were included in the multivariate analysis, and a p-value of <0.05 in the multivariable analysis was considered statistically significant.

Weighting adjustments were performed to account for the non-random elements of the Respondent-Driven Sampling technique (RDS). Composite scores for participant responses in the Health Belief Model (HBM) constructs were calculated. A score below the average for each specific construct is considered low.

### Ethical considerations

This study observed all ethical procedures. Ethical approval was obtained from Kilimanjaro Christian Medical University College (PG02/2024), the principal investigator’s institution. Approval was also obtained from all local ethics committees to ensure the study adhered to regional and institutional guidelines. All participants were anonymously identified to maintain confidentiality and privacy.

## Results

The average age of participants was (36.11±5.243 SD) years, and almost half, 173(49.3%), were between 25-34 years. More than half, 232(66.1%), and 183(52.3%) resided in urban areas and completed primary education, respectively. Almost half, 175(49.9%), were separated or divorced, and seventy per cent, 144(69.5%), had two or more children. The majority, 279(75.9%), had tested negative for HIV, and almost eighty per cent, 280(79.8%), reported using modern contraceptives. (Table 1)

**Table 1:** Sociodemographic and behavioural characteristics of FSWs in Kilimanjaro region.

| Variable | Frequency | Percentage |
| --- | --- | --- |
| **Age (Years), Mean ± SD** | 36.11±5.243 | |
| 25-34 | 173 | 49.3 |
| 35-39 | 91 | 25.9 |
| 40-44 | 62 | 17.7 |
| 45-49 | 25 | 7.1 |
| **Residence** |  |  |
| Rural | 119 | 33.9 |
| Urban | 232 | 66.1 |
| **Education** |  |  |
| Never went to school | 13 | 3.7 |
| Primary | 183 | 52.1 |
| Secondary | 146 | 41.6 |
| College/University | 9 | 2.6 |
| **Marital Status** |  |  |
| Single | 111 | 31.6 |
| Married/Cohabiting | 65 | 18.5 |
| Separated/Divorced | 175 | 49.9 |
| **Parity** |  |  |
| Nulliparous/Para 1 | 107 | 30.5 |
| Para 2+ | 244 | 69.5 |
| **HIV status** |  |  |
| Positive | 35 | 10.0 |
| Negative | 279 | 79.5 |
| Unknown | 37 | 10.5 |
| **Use contraceptives** |  |  |
| Yes | 280 | 79.8 |
| No | 71 | 20.2 |
| **Knowledge of cervical cancer** |  |  |
| High | 70 | 19.9 |
| Low | 281 | 80.1 |
| **Perceived susceptibility** |  |  |
| High | 27 | 7.7 |
| Low | 324 | 92.3 |
| **Perceived Severity** |  |  |
| High | 115 | 32.8 |
| Low | 236 | 67.2 |
| **Perceived Benefits** |  |  |
| High | 143 | 40.7 |
| Low | 208 | 59.3 |
| **Perceived Barriers** |  |  |
| High | 40 | 11.4 |
| Low | 311 | 88.6 |
| **Self-Efficacy** |  |  |
| High | 99 | 28.2 |
| Low | 252 | 71.8 |
| **Cues to Action** |  |  |
| High | 111 | 31.6 |
| Low | 240 | 68.4 |

Based on responses to cervical cancer and screening from the HBM perspective, the majority, 324(92.3%) of participants, reported being less susceptible to cervical cancer, and more than half, 236(67.2%) had less perception of the severity of cervical cancer disease. On cervical cancer screening, more than one-sixth, 208(59.3%) of participants perceived cervical cancer screening as being beneficial to them. The majority, 311(88.6%) of participants perceived fewer barriers to navigating cervical cancer screening services. Almost seventy-two per cent, 252(71.8%), reported having low self-efficacy for utilising cervical cancer screening services, and more than half, 240(68.4%), reported being motivated by different cues. (Table 1)

The perceptions corresponded with the mean scores: perceived susceptibility was relatively low at 2.66. In contrast, perceived severity had a higher mean score of 3.27. while perceived benefits were 3.40. Perceived barriers had a moderate mean score of 2.91. Also, participants exhibited a relatively high sense of self-efficacy, with a mean score of 3.37, while motivational factors, represented by cues to action, had a mean score of 3.21(Table 3). Regarding cervical cancer screening uptake, only 17(4.8%) had ever been screened with younger age groups having a lower screening rate 6(3.5%), compared to older age groups.

There was a difference in screening rates among FSWs from urban compared to their rural counterparts 13(5.6%) vs 4(3.4%). Those with a college or university education had a bit higher screening rate 1(11.1%) compared to other levels (Table 2).

**Table 2:** Sociodemographic Characteristics, HBM Constructs, and Cervical Cancer Screening Uptake among FSWs in Kilimanjaro region (N=351)

| Variable | Ever been screened for cervical cancer? n (%) |  | P-Value |
| --- | --- | --- | --- |
|  | Yes | No |  |
| **Age (Years)** |  |  |  |
| 25-34 | 6(3.50) | 167(96.5) | 0.147 |
| 35-39 | 3(3.30) | 88(96.70) |  |
| 40-44 | 5(8.10) | 57(91.90) |  |
| 45-49 | 3(12.5) | 21(87.50) |  |
| **Residence** |  |  |  |
| Rural | 4(3.40) | 115(96.6) | 0.354 |
| Urban | 13(5.6) | 219(94.4) |  |
| **Education** |  |  |  |
| Never went to school | 1(7.70) | 12(92.30) | 0.781 |
| Primary | 8(4.40) | 175(95.6) |  |
| Secondary | 7(4.80) | 139(95.2) |  |
| College/University | 1(11.10) | 8(88.90) |  |
| **Marital Status** |  |  |  |
| Single | 4(3.60) | 107(96.4) | 0.724 |
| Married/Cohabiting | 4(6.20) | 61(93.80) |  |
| Separated/Divorced | 9(5.10) | 166(94.9) |  |
| **Parity** |  |  |  |
| Nulliparous/Para 1 | 1(0.90) | 106(99.1) | **0.024** |
| Para 2+ | 16(6.60) | 228(93.4) |  |
| **HIV status** |  |  |  |
| Positive | 0(0.00) | 35(100.0) | 0.100 |
| Negative | 17(6.10) | 262(93.9) |  |
| Unknown | 0(0.00) | 37(100.0) |  |
| **Use contraceptives** |  |  |  |
| Yes | 10(3.60) | 270(96.4) | **0.028** |
| No | 7(9.90) | 64(90.10) |  |
| **Knowledge of cervical cancer** |  |  |  |
| High | 4(5.70) | 66(94.30) | 0.704 |
| Low | 13(4.60) | 268(95.4) |  |
| **Perceived susceptibility** |  |  |  |
| High | 2(7.40) | 25(92.60) | 0.518 |
| Low | 15(4.60) | 309(95.4) |  |
| **Perceived Severity** |  |  |  |
| High | 10(8.70) | 105(91.3) | **0.019** |
| Low | 7(3.00) | 229(97.0) |  |
| **Perceived Benefits** |  |  |  |
| High | 12(8.40) | 131(91.6) | **0.010** |
| Low | 5(2.40) | 203(97.6) |  |
| **Perceived Barriers** |  |  |  |
| High | 3(7.50) | 37(92.50) | 0.406 |
| Low | 14(4.50) | 297(95.5) |  |
| **Self-Efficacy** |  |  |  |
| High | 10(10.1) | 89(89.90) | **0.004** |
| Low | 7(2.80) | 245(97.2) |  |
| **Cues to Action** |  |  |  |
| High | 10(9.0) | 101(91.0) | **0.013** |
| Low | 7(2.90) | 233(97.1) |  |

KEY: **HBM**: Health Belief Model

**Table 3:** Descriptive analysis of participants’ response rates on Health Belief Model Constructs item questions regarding cervical cancer screening uptake in Kilimanjaro region (N=351)

| **HBM Constructs** | **SD (1)** | **D (2)** | **N (3)** | **A (4)** | **SA (5)** | **Mean ±SD** |
| --- | --- | --- | --- | --- | --- | --- |
| **Perceived Susceptibility to Cervical Cancer** | **Cronbach's Alpha= 0.76** |  |  |  |  |  |
| I feel like I will get cervical cancer sometime in my life | 57(16.2) | 92(26.2) | 158(45.0) | 15(4.3) | 29(8.3) | 2.62±1.070 |
| If I continue with this lifestyle, cervical cancer is likely | 48(13.7) | 93(26.5) | 159(45.3) | 29(8.3) | 22(6.3) | 2.67±1.019 |
| I feel the probability of getting cervical cancer in a few years is high to me. | 54(15.4) | 101(28.8) | 135(38.5) | 27(7.7) | 34(9.7) | 2.68±1.125 |
| **Total mean score** |  |  |  |  |  | **2.66** |
| **Perceived Severity of Cervical Cancer** | **Cronbach's Alpha 0.87** |  |  |  |  |  |
| I'm afraid of thinking about this disease. | 38(10.8) | 68(19.4) | 115(32.8) | 45(12.8) | 85(24.2) | 3.20±1.299 |
| The thought of this disease scares me a | 36(10.3) | 71(20.2) | 102(29.1) | 46(13.1) | 96(27.4) | 3.27±1.330 |
| When I think about cervical cancer, my heart beats faster | 23(6.6) | 65(18.5) | 115(32.8) | 55(15.7) | 93(26.5) | 3.37±1.237 |
| If I had this disease, my whole life would change | 25(7.1) | 78(22.2) | 101(28.8) | 56(16.0) | 91(25.9) | 3.31±1.269 |
| Problems I would experience with this disease would last a long time | 21(6.0) | 89(25.4) | 113(32.2) | 41(11.7) | 87(24.8) | 3.24±1.244 |
| This disease would threaten my relationship | 28(8.0) | 75(21.4) | 125(35.6) | 45(12.8) | 78(22.2) | 3.20±1.231 |
| **Total mean score** |  |  |  |  |  | **3.27** |
| **Perceived Benefits of CC Screening** | **Cronbach's Alpha= 0.92** |  |  |  |  |  |
| Sustaining good health is very important to me | 41(11.7) | 59(16.8) | 93(26.5) | 49(14.0) | 109(31.1) | 3.36±1.376 |
| Having consistent screening helps me detect changes before they become cancer | 33(9.4) | 50(14.2) | 110(31.3) | 47(13.4) | 111(31.6) | 3.44±1.316 |
| I want to realise health complications early | 31(8.8) | 56(16.0) | 105(29.9) | 49(14.0) | 110(31.3) | 3.43±1.314 |
| I think it is vital to do things that improve health | 33(9.4) | 48(13.7) | 106(30.2) | 61(17.4) | 103(29.3) | 3.44±1.294 |
| I think regular screening is the best way | 32(9.1) | 51(14.5) | 101(28.8) | 58(16.5) | 109(31.1) | 3.46±1.308 |
| I think finding the right information is important | 23(6.6) | 59(16.8) | 111(31.6) | 55(15.7) | 103(29.3) | 3.44±1.252 |
| I believe regular screening lowers the risk of this disease | 26(7.4) | 67(19.1) | 104(29.6) | 58(16.5) | 96(27.4) | 3.37±1.270 |
| I think early detection improves treatment outcomes. | 29(8.3) | 66(18.8) | 118(33.6) | 55(15.7) | 83(23.6) | 3.28±1.245 |
| **Total mean score** |  |  |  |  |  | **3.40** |
| **Perceived Barriers** | **Cronbach's Alpha 0.75** |  |  |  |  |  |
| I'm afraid because I don't know what will happen | 36(10.3) | 56(16.0) | 138(39.3) | 68(19.4) | 53(15.1) | 3.13±1.164 |
| I don't like to hear about the bad result | 36(10.3) | 67(19.1) | 134(38.2) | 67(13.4) | 47(13.4) | 3.06±1.152 |
| I feel embarrassed to be examined | 39(11.1) | 83(23.6) | 134(38.2) | 66(18.8) | 29(8.3) | 2.89±1.092 |
| My partner won't allow me to get screened | 62(17.7) | 87(24.8) | 129(36.8) | 52(14.8) | 21(6.0) | 2.67±1.111 |
| I won't get an appointment for screening | 51(14.5) | 74(21.1) | 144(41.0) | 58(16.5) | 24(6.8) | 2.80±1.093 |
| I won't go for the screening if I have to pay for it | 49(14.0) | 79(22.5) | 123(35.0) | 64(18.2) | 36(10.3) | 2.88±1.169 |
| This disease may affect the intrauterine devices | 35(10.0) | 84(23.9) | 136(38.7) | 63(17.9) | 33(9.4) | 2.93±1.092 |
| I won't get time to go for cervical cancer screening | 42(12.0) | 78(22.2) | 136(38.7) | 57(16.2) | 38(10.8) | 2.92±1.137 |
| **Total mean score** |  |  |  |  |  | **2.91** |
| **Self-Efficacy** | **Cronbach's Alpha 0.81** |  |  |  |  |  |
| I believe I'm capable of going for it | 36(10.3) | 70(19.9) | 0(0) | 198(56.4) | 47(13.4) | 3.43±1.237 |
| I believe I can tolerate the distress of screening | 35(10.0) | 57(16.2) | 11(3.1) | 194(55.3) | 54(15.4) | 3.50±1.219 |
| I believe I can arrange a time and go for the screening | 22(6.3) | 69(19.7) | 1(0.3) | 189(53.8) | 70(19.9) | 3.62±1.187 |
| I believe that exercising regularly lowers the risk of this | 31(8.8) | 60(17.1) | 143(40.7) | 61(17.4) | 56(16.0) | 3.15±1.148 |
| I believe, having regular check-ups can help | 23(6.6) | 71(20.2) | 132(37.6) | 74(21.1) | 51(14.5) | 3.17±1.110 |
| **Total mean score** |  |  |  |  |  | **3.37** |
| **Cues to Action** | **Cronbach's Alpha 0.75** |  |  |  |  |  |
| The campaigns would motivate me | 23(6.6) | 72(20.5) | 129(36.8) | 90(25.6) | 37(10.5) | 3.13±1.064 |
| If I get advice from my friends, I will go | 19(5.4) | 64(18.2) | 152(43.3) | 86(24.5) | 30(8.5) | 3.13±0.986 |
| I like to be examined by a female provider | 21(6.0) | 59(16.8) | 140(39.9) | 89(25.4) | 42(12.0) | 3.21±1.049 |
| If no cost, then I will go for it | 14(4.0) | 55(15.7) | 137(39.0) | 105(29.9) | 40(11.4) | 3.29±0.995 |
| If health professionals recommend it, I will do it | 18(5.1) | 55(15.7) | 125(35.6) | 118(33.6) | 35(10.0) | 3.28±1.012 |
| **Total mean score** |  |  |  |  |  | **3.21** |

**KEY:** CC: Cervical Cancer; SD: Strongly Disagree; D: Disagree; N: Neutral; A: Agree; SA: Strongly Agree; ± SD: Standard Deviation; HBM: Health Belief Model; FSWs: Female Sex Workers.

Among behavioural factors influencing cervical cancer screening uptake from the HBM perspective, four factors; Perceived severity(AOR: 3.25, 95%CI:1.16-9.07), perceived benefits (AOR:3.61,95% CI:1.10-11.84), self-efficacy (AOR: 3.59, 95% CI: 1.18-10.96) and cues to action (AOR: 3.61, 95% CI: 1.28-10.15) were positively associated with cervical cancer screening uptake among this population. However, this study found no significant association between perceived susceptibility (AOR: 1.65, 95% CI: 0.36-7.62), perceived barriers (AOR: 1.72, 95% CI: 0.47-6.27) and screening uptake. (Table 4).

**Table 4:** Binary logistic analysis of behavioural determinants of Cervical Cancer Screening Uptake among FSWs in Kilimanjaro. (N=351)

| Variable | <u>Screening Uptake</u><br>n (%) |  | <u>Bivariate Analysis</u><br>COR (95% CI) | P-value | <u>Multivariate Analysis</u><br>AOR (95% CI) | P-value |
| --- | --- | --- | --- | --- | --- | --- |
|  | Yes | No |  |  |  |  |
| **Perceived susceptibility** |  |  |  |  |  |  |
| High | 2(7.40) | 25(92.60) | 1.65(0.36-7.62) | 0.522 |  |  |
| Low | 15(4.60) | 309(95.4) | 1 |  |  |  |
| **Perceived Severity** |  |  |  |  |  |  |
| High | 10(8.70) | 105(91.3) | 3.12(1.15-8.41) | **0.025** | 3.25(1.16-9.07) | **0.025** |
| Low | 7(3.00) | 229(97.0) | 1 |  | 1 |  |
| **Perceived Benefits** |  |  |  |  |  |  |
| High | 12(8.40) | 131(91.6) | 3.72(1.28-10.81) | **0.016** | 3.61(1.10-11.84) | **0.034** |
| Low | 5(2.40) | 203(97.6) | 1 |  | 1 |  |
| **Perceived Barriers** |  |  |  |  |  |  |
| High | 3(7.50) | 37(92.50) | 1.72(0.47-6.27) | 0.411 |  |  |
| Low | 14(4.50) | 297(95.5) | 1 |  |  |  |
| **Self-Efficacy** |  |  |  |  |  |  |
| High | 10(10.1) | 89(89.90) | 3.93(1.45-10.65) | **0.007** | 3.59(1.18-10.96) | **0.024** |
| Low | 7(2.80) | 245(97.2) | 1 |  | 1 |  |
| **Cues to Action** |  |  |  |  |  |  |
| High | 10(9.0) | 101(91.0) | 3.29(3.22-8.90) | **0.019** | 3.61(1.28-10.15) | **0.015** |
| Low | 7(2.90) | 233(97.1) | 1 |  | 1 |  |

KEY: **COR**: Crude Odds Ratio, **AOR**: Adjusted Odds Ratio; **CI**: Confidence Interval; **FSWs**: Female sex workers

## DISCUSSION

The uptake of cervical cancer screening was significantly low among FSWs in the Kilimanjaro region, given their increased vulnerability to cervical cancer. This low uptake of cervical cancer screening among this population may be attributed to the low perception of susceptibility to cervical cancer. This finding aligns with the findings from studies conducted in Bangladesh and Nigeria, where similar low screening rates among FSWs were reported. [14].

In contrast, these findings differ from studies conducted in Ethiopia and Kenya, where much higher cervical cancer screening uptake among FSWs was reported. [15], [16]. This could be due to the presence of targeted interventions tailored to the needs of FSWs in those regions as compared to our setting. The results of the current study imply that well-designed programs may significantly reduce both perceptual and structural barriers, leading to improved screening uptake.

This study found a significant positive influence of perceived severity, perceived benefits, self-efficacy, and cues to action on the uptake of cervical cancer screening among this population. These observations are in line with the HBM constructs which postulate that when an individual (i.e., FSWs) recognizes the serious consequences of cervical cancer (i.e., perceived severity), and understand the advantages of screening (i.e., perceived benefits), participants are more likely to engage in cervical cancer screening services. [17] Additionally, these findings reveal that when FSWs believe in their ability to undergo screening (i.e., Self-efficacy), and the presence of reminders or encouragement from healthcare providers (i.e., Cues to actions), can motivate FSWs, and are more likely to engage in cervical cancer screening services. These findings are consistent with several existing studies conducted in different regions.

For instance, our results align with a study conducted in South Africa, which reported a significant positive association between perceived severity and cervical cancer screening uptake among FSWs [18]. Similarly, a study these findings align with findings from a study done in South India state, Karnataka, and Ethiopia[19], [20]. This similarity may be attributed to the shared high-risk sexual behaviours among FSWs in both contexts.

In contrast, our findings differ from a study in Jordan, which found no association between perceived severity and screening uptake. This discrepancy suggests that cultural context may influence how risk perceptions affect health behaviours, such as cervical cancer screening, and underscores the importance of culturally tailored interventions.[21]

Regarding, perceived benefits, our findings align with studies conducted in Ethiopia, which revealed similar results[20], [22]. This similarity can be explained by increased awareness of the availability of cervical cancer screening services and the corresponding benefits in both regions, which might play a role in motivating FSWs to participate in screening services.

Regarding Self-efficacy as a strong predictor of cervical cancer screening uptake our findings are consistent with the findings from a similar study done in Ethiopia which revealed a strong positive association between self-efficacy and cervical cancer screening uptake among FSWs[22]. This similarity can be explained by a similar motivational role in behaviour change among FSWs. Conversely, a study done in Iran revealed a weaker positive association between self-efficacy and cervical cancer screening uptake among FSWs, suggesting that self-confidence does not necessarily play a significant role in motivating FSWs to utilize cervical cancer screening services. This difference can be explained by the variations in individual perceptions as a result of different cultural contexts [23].

Regarding cues to action as a predictor for uptake of cervical cancer screening among FSWs, our findings were consistent with the previous findings reported in a systematic review done in Ethiopia, which also revealed a strong significant association between cues to action and cervical cancer screening uptake among FSWs [24]. This suggests that certain triggers or prompts significantly motivate FSWs to utilize cervical cancer screening services [24]. In contrast, the findings were different from a study done in Japan, which revealed no association between cues to action and cervical cancer screening uptake among FSWs [25]. This difference suggests that differences in cue relevance and delivery methods may drive variability across settings.

## Conclusion

The uptake of cervical cancer screening among FSWs in Kilimanjaro region was significantly low despite high levels of awareness. Targeted interventions tailored to the specific needs of this population are essential. Much focus should be directed toward addressing key behavioural determinants: increasing the perceived severity of cervical cancer, emphasizing the benefits of screening, enhancing self-efficacy, and maximizing the effectiveness of cues to action.

## Acknowledgment

Our sincere appreciation should go to all the participants who generously shared their time and experiences. Special thanks to KCRI, KCMUCo, and TAWREF for their invaluable support and collaboration throughout this study.

## Author Contribution

G. D. S: Conceptualized the study, proposal writing, data collection, data analysis, interpretation, report writing, and manuscript preparation.

I. H. U: Reviewed the manuscript for clarity and rigor.

B. N.: Conceptualization of the theoretical framework.

E. M: Involved in proposal writing.

P. S: Reviewed the manuscript for clarity and rigor.

A.R: Data collection

G.O: Data collection

J.E. M: Reviewed the manuscript for clarity and rigor.

P.M: Data collection

L. M: Data collection

H.K: Data collection

B.M: Study conceptualization

B.T. M: Study conceptualization, approved funding, and guidance to enhance the study’s quality.

A.M: Study Conceptualization, Supervised all stages, Critical review of the manuscript.

## Patient consent for publication

All participants in this study gave consent.

## Data availability statement

The dataset for this work is available upon request to the corresponding author.

## Open access

This is an open-access article provided there is a proper citation.

## Declaration of AI Assistance

We hereby declare that AI technology (ChatGPT) was used solely to improve the readability and language of this work, without modifying the core ideas or content. All research tasks, data analysis, findings, and interpretations presented in this document are entirely based on our original data and were independently conducted by us.

